# Identifying and replicating plasma proteins associated with hypertrophic cardiomyopathy severity in carriers of pathogenic *MYBPC3* variants

**DOI:** 10.64898/2026.03.28.26349616

**Authors:** Fahima Hassanzada, Marion van Vugt, Mark Jansen, Annette F. Baas, Anneline S.J.M. te Riele, Dennis Dooijes, Saskia N. van der Crabben, Jan D.H. Jongbloed, Moniek G.P.J. Cox, Ahmad S. Amin, Ronald H. Lekanne Deprez, Bram Ruijsink, Diederik W. D. Kuster, Jolanda van der Velden, Connie R. Bezzina, Folkert W. Asselbergs, J. Peter van Tintelen, Karin Y. van Spaendonck-Zwarts, A. Floriaan Schmidt

## Abstract

**Background:** Hypertrophic cardiomyopathy (HCM) is a clinically variable disease in terms of onset and progression. Pathogenic *MYBPC3* variants account for a substantial proportion of HCM diagnoses. This study sought to identify protein biomarkers associated with HCM severity.

**Methods:** Olink-assayed plasma proteins of 144 *MYBPC3* pathogenic variant carriers were tested for associations with HCM severity based on HCM diagnostic criteria (unaffected, mildly, or severely affected). The UK Biobank was used to replicate the identified proteins through considering time to onset of HCM (67 cases), cardiomyopathy (156 cases),and associations with cardiac MRI derived left ventricular maximum wall thickness (6,492 participants). Replicated proteins were further prioritised based on cardiac tissue expression and druggability, and annotated using pathway enrichment and association with onset of: heart failure (HF), dilated cardiomyopathy (DCM), sudden cardiac arrest (SCA), and ventricular arrhythmias (VA).

**Results:** Among pathogenic *MYBPC3* variant carriers, we identified 27 proteins associated with HCM severity. We independently replicated 21 proteins in the UK Biobank. Of the five prioritised proteins (NT-proBNP, GDF-15, FGF-23, ADM, and NCAM1), all but NT-proBNP were targeted by drugs with repurposing potential. The replicated proteins additionally associated with the incidence of HF (n=5), DCM (n=4), SCA (n=4), and VA (n=4).

**Conclusion:** This study replicated 21 and prioritised five proteins associated with HCM severity in pathogenic *MYBPC3* variant carriers. Replication in unselected HCM suggests the prioritised proteins are associated with HCM independent of genotype, providing important leads for plasma-based markers for diagnoses, disease monitoring, and drug targets.

## Introduction

Hypertrophic cardiomyopathy (HCM) is defined as left ventricular hypertrophy not explained by abnormal loading conditions^1,2^. In up to 40% of HCM patients, a rare pathogenic genetic variant can be identified^2^ and approximately 40% of these harbor a variant in *MYBPC3*^3^. HCM may present with left ventricular (LV) outflow tract obstruction, and can lead to complications such as sudden cardiac arrest (SCA) or congestive heart failure (HF)^4,5^. Conversely, many patients with HCM exhibit mild symptoms or remain asymptomatic and penetrance among pathogenic variant carriers is incomplete^6^.

The recently developed cardiac myosin inhibitors have demonstrated the feasibility of targeted drug development for HCM^7^. Given the variable severity of HCM and its incomplete penetrance, it is crucial to identify additional drug targets for HCM complementing myosin inhibitors as well as identify biomarkers relevant for (early) disease detection and monitoring. Proteomic markers from plasma are especially suitable for this, as they may reflect key biological, potentially druggable effector pathways, and can be readily measured, monitored, and targeted by drugs.

In the current study, we aimed to identify plasma proteins related with HCM severity sourcing Olink plasma proteomics measurements from the BIO FOr CARe study^8^. This study enrolled individuals carrying pathogenic *MYBPC3* variants implicated with mRNA truncation and haploinsufficiency, impairing cardiomyocyte function^9–11^. Proteins associating with HCM severity were subsequently replicated using Olink protein measurements available from up to 52,560 UK Biobank (UKB) participants, determining associations with onset of HCM, any cardiomyopathy (CMP), and cardiac MRI (CMR) derived LV maximum wall thickness (LVMWT). Considering cardiac tissue mRNA expression and protein druggability, five replicated proteins were prioritised (NT-proBNP, GDF-15, FGF-23, ADM, and NCAM1).

These prioritised proteins were additionally associated with onset of HF, and all but NCAM1 also with dilated cardiomyopathy (DCM), SCA, or ventricular arrhythmia (VA). Four of these prioritised proteins were targeted by an approved drug or drug tested in clinical trials, providing opportunities for drug repurposing and development.

## Methods

### Discovery and replication cohort studies

The BIO FOr CARe (Identification of BIOmarkers of hypertrophic cardiomyopathy development and progression in Dutch MYBPC3 FOunder variant CARriers) study was initiated to identify plasma proteins associating with HCM onset and severity. The study enrolled HCM patients and their family members carrying at least one of the pathogenic *MYBPC3* founder variants: NM_000256.3 c.2373dupG p.(Trp792Valfs*41), c.2827C>T p.(Arg943*), c.2864_2865delCT p.(Pro955Argfs*95), or c.3776delA p.(Gln1259Argfs*72). Participants under 18 years of age and those with prior heart transplantation were excluded. Inclusion started in January 2017 and continued until June 2024^11^. This study was performed in accordance with the Helsinki declaration and was approved by the Medical Ethics Committee of the UMC Utrecht under registration number NL5588904115. Written informed consent was obtained from all participants. The *MYBPC3* carriers were classified as unaffected, mildly affected, or severely affected. Unaffected carriers did not meet HCM diagnostic criteria, whereas mildly affected patients did meet the criteria but did not have any of the following criteria: LVMWT ≥ 20mm, an LV outflow tract obstruction necessitating septal reduction therapy, HF diagnosis, or malignant VA. Severely affected patients had at least one of the aforementioned criteria. Measurements and diagnoses were sourced from available electronic healthcare records data using a window of one year before and one year after study enrolment, taking the measurement closest to enrolment.

The subset of proteins associating with HCM severity in the BIO FOr CARe study were replicated by determining their associations with time to onset of HCM, CMP, and cross-sectionally with LVMWT (in mm) in UK Biobank (UKB) participants free of any CMP at the time of Olink plasma protein measurement. Proteomics measurements were available in 52,560 participants, of whom 6,942 also had LVMWT data from CMR performed in the first wave of the CMR sub-study, please see the protocol as detailed elsewhere^12^. Briefly, all CMR examinations were performed on a 1.5 Tesla scanner (Magnetom Aera, Syngo Platform VD13A, Siemens Healthcare, Erlangen, Germany). We used a previously developed and validated deep-learning methodology (AI-CMR^QC^) to extract LV CMR measurements^13^ and cine images of short-axis and 2- and 4-chamber long-axis views were used to automatically calculate the LVMWT. Disease diagnoses of HCM and CMP were sourced from integrated electronic healthcare records from general practitioners, hospital episode statistics, and death registries (**Appendix Table S1-2**).

### Proteomics

In BIO FOr CARe, targeted proteomics was performed at the Central Diagnostic Laboratory of the University Medical Center Utrecht using the Olink Target 96 Cardiovascular II, Cardiovascular III, and Cardiometabolic panels, each assessing 92 unique proteins. Protein measurements were normalised to control samples and expressed on a log_2_ scale. Following standard guidance from Olink, we removed assays where 25% or more of the samples had a value below the limit of detection. Proteins were further pruned on low variability (threshold of 0.05) and pairwise correlation of 0.90 or higher, removing BNP and LOX-1.

The UKB pharma proteomics project measured 2,923 protein analytes across the Olink panels Oncology, Oncology II, Neurology, Neurology II, Inflammation, Inflammation II, Cardiometabolic, and Cardiometabolic II. Detailed information on processing and quality control is described elsewhere^14^. Similar to the BIO FOr CARe study, proteins where more than 25% of the samples had a value below the limit of detection were excluded. After quality control, only one protein (MMP-2) measured in BIO FOR CARe was not available in the UK Biobank.

### Statistical analysis

To identify proteins associated with HCM severity in the BIO FOr CARe study, we performed two random forest analyses on all proteins jointly: 1) focussing on identifying proteins associating with a specific severity class using a one-vs-one contrast (referred to as HCM severity comparisons), 2) comparing all affected to unaffected participants. Random forest was chosen to flexibly account for potential non-linear associations. Hyper-parameter tuning was performed using a randomised grid-search, where a random subset of 50 hyper-parameter combinations was evaluated using fivefold cross-validation. Optimisation was based on the (macro) F1-score, and considered the following hyper-parameters: minimum samples per terminal leaf, minimum samples required to split a node, maximum considered features per tree, number of trees, and maximum tree depth. Given that a random forest algorithm only uses a random subset of samples, the hold-out sample was used to estimate the out-of-bag performance in terms of outcome-specific F1-score. Feature importance was determined using the Gini index, where a value of 1 indicates a terminal leaf without class variability (i.e., everyone in the terminal leaf experiences the same outcome).

We identified the top 10 most important proteins for each HCM severity comparison and sought to externally validate these in the UKB by considering association with incident HCM, incident CMP, and LVMWT in individuals free of CMP at protein measurement. The associations with HCM and CMP were derived from a Cox proportional hazards model adjusted for age and sex. Participants where censored when lost-to-follow-up, died, or end of study, whichever occurred first. Linear regression was performed to determine the associations with LVMWT, again adjusted for age and sex. In both datasets missing protein values were imputed using sklearn’s K-nearest neighbours algorithm (using k=5).

Results are presented as hazard ratios (HR) or as mean difference, accompanied by 95% confidence intervals (CI). In the replication analyses we employed an alpha of 0.05 for nominal significance, and a multiplicity corrected alpha of 1.85×10^-3^, based on a Bonferroni-adjustment for 27 proteins.

### Protein annotation and prioritisation

Proteins were annotated based on tissue expression, druggability, and pathway analysis. We sourced tissue mRNA expression data from the human protein atlas v20^15^ and compared the cardiac mRNA expression with the mean expression across the other available tissues to identify proteins that were overexpressed in the heart compared to other tissues. Pathway analysis was performed by querying the Reactome v91 knowledgebase^16^ to map all Olink-measured proteins to biological pathways. Enrichment was assessed by determining the difference between the proportion of prioritised proteins involved in each pathway and the proportion of all the available Olink-measured proteins involved in the same pathway, this difference was evaluated using a Wald-based test. For druggability, we obtained information on indications and side-effects from ChEMBL v33 and considered proteins as “drugged” if targeted by an approved compound and “druggable” if targeted by a compound in development.

The proteins were assigned a prioritisation score based on discovery (BIO FOr CARe), replication (UKB), and annotation. One point was assigned for each HCM severity comparison (e.g. mild vs. unaffected, severe vs. unaffected) in which the protein ranked among the top ten selected features, thereby giving higher scores to proteins consistently affecting multiple severity stages. An association with each of the replication outcomes (HCM, CMP, and LVMWT) was similarly assigned a single point, with further points assigned for overexpression in cardiac tissue and protein druggability (one point for potentially druggable and two points for a drugged protein targeted by an approved compound). Proteins with a summed score of five points or more, including at least one replication point, were prioritised.

The prioritised proteins were further annotated by considering associations with HCM-relevant outcomes: HF, DCM, VA, and SCA in the UKB, reflecting classical HCM endpoints (HF, VA, and SCA) and offering insight into molecular pathways of ventricular remodeling (DCM); see **Appendix Table S1-2**. As before, an alpha of 0.05 was considered nominally significance, with a Bonferroni-adjusted alpha of 2.50×10^-3^ accounting for the number of tests.

## Results

### Identification of proteins involved in HCM onset and severity in MYBPC3 variant carriers

Random Forests were applied to 268 plasma proteins measured in 144 *MYBPC3* variant carriers categorised as unaffected (n=52, 75% female, median LVMWT 10.0 mm [Q1 9.0; Q3 10.6]), mildly affected (n=17, 59% female, median LVMWT 15.0 mm [Q1 13.1; Q3 16.0]), and severely affected (n=75, 31% female, median LVMWT 18.0 mm [Q1 15.2; Q3 21.8]; **Appendix Table S3**). Selecting the ten features with the highest importance per HCM severity comparisons (i.e. unaffected vs. affected, unaffected vs. mildly affected, unaffected vs. severely affected, mildly vs. severely affected) identified 27 proteins (**Figure 1-2**, **Appendix Table S4**). F1 scores are listed in **Appendix Table S5**. Three proteins (NT-proBNP, GDF-15, and ACE2) were among the top ten for all HCM severity comparisons indicating HCM onset. Cystatin C and NCAM1 were important when comparing affected and mildly affected to unaffected. MERTK, ADM, REN, and FGF-23 were important when comparing affected and severely to unaffected participants, while SCF was important when comparing unaffected and mildly affected with severely affected participants; **Appendix Figure S1**).

**Figure 1.**
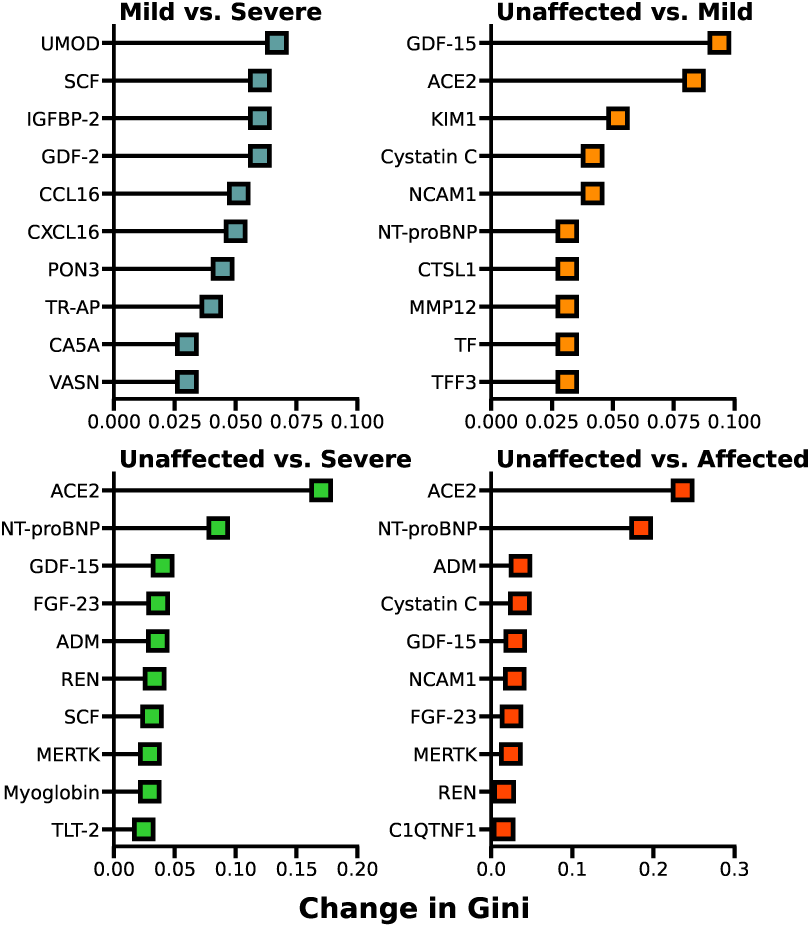
Top ten most important proteins per HCM severity comparisons. Random Forests were applied to 268 plasma proteins measured in 52 unaffected, 17 mildly affected, and 75 severely affected *MYBPC3* variant carriers. Feature importance is ranked by the Gini index, where a value of 1 indicates a terminal leaf without class variability.

**Figure 2.**
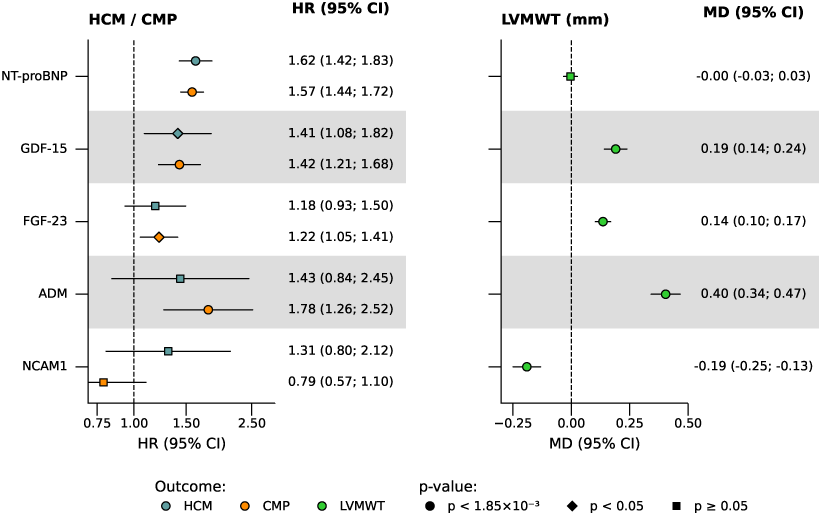
Overview of the prioritisation strategy of proteins associating with HCM severity. Each column indicates the number of points assigned to each protein based on the association or annotation of the protein. Specifically, the first column indicates the number of HCM severity comparisons (i.e. unaffected versus affected, unaffected versus mild, unaffected versus severe, and mild versus severe) in which the protein was among the top ten selected features. The second and third column indicates whether the protein was replicated in the UKB. The fourth column indicates the extent of cardiac expression and the fifth column the druggability. The last column is a sum of the previous column and indicates the prioritisation score. Abbreviations: CMP = cardiomyopathy; Disc = discovery; HCM = hypertrophic cardiomyopathy; LVMWT = left ventricular maximum wall thickness.

### Prioritisation of HCM related proteins

To prioritise the proteins associated with HCM severity, we assessed their association with time to incident HCM (67 cases),CMP (156 cases), and the association with LVMWT (n=6,492). Twenty-one proteins were associated with at least one of these outcomes (**Figure 2**, **Appendix Table S6**). Selecting the proteins with a prioritisation score of five or more, we identified a subset of five replicated proteins with strong evidence for involvement with HCM phenotype: NT-proBNP, GDF-15, FGF-23, ADM, and NCAM1. GDF-15 (nominally) associated with increased risk of HCM and CMP, as well as with larger LVMWT (**Figure 3**). NT-pro-BNP associated with both HCM and CMP (HR 1.62; 95%CI 1.42; 1.83 for HCM, and HR 1.57; 95%CI 1.44; 1.72 for CMP), while FGF-23 and ADM associated with higher LVMWT and either increased HCM or CMP risk (**Figure 2-3**).

**Figure 3.**
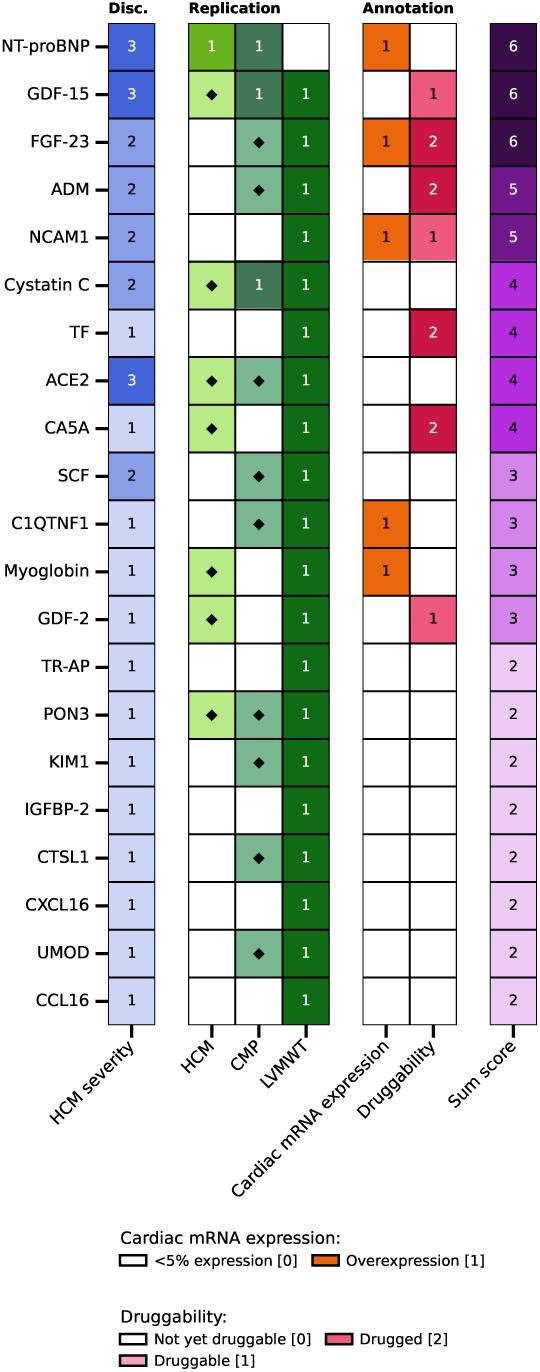
Effect estimates for the replicated proteins associating with HCM, CMP, or LVMWT. Proteins were considered significantly replicated if the p-value was below the p-value threshold of 0.05 divided by the 27 for number of proteins tested. Abbreviations: CI = confidence interval; CMP = cardiomyopathy; HCM = hypertrophic cardiomyopathy; LVMWT = left ventricular maximum wall thickness; MD = mean difference.

### Annotation of proteins associated with HCM

We next explored the biological function of the prioritised proteins. While all five prioritised proteins were expressed in cardiac tissue, NT-proBNP, FGF-23, and NCAM1 were overexpressed in heart tissue compared to non-heart tissues (**Appendix Figure S2**, **Appendix Table S7**). Considering pathway enrichment, we found that ADM is involved in G alpha (s) subunit signalling, which plays a role in cAMP production. FGF-23 and NCAM1 are both involved in the Ras-MAPK pathway but this was not sufficient for statistical enrichment (p=0.091; **Appendix Figure S3**, **Appendix Table S8**). Linking the prioritised proteins to information on drug compounds showed that ADM and FGF-23 are affected by approved drugs: ADM is targeted by enibarcimab, an antibody in development for HF, while FGF-23 is inhibited by burosumab to treat bone diseases (including X-linked hypophosphatemia).

Furthermore, GDF-15 is targeted by the phase-2 compounds visugromab and ponsegromab, which are being tested for HF and cancer, including cachexia, while NCAM1 antibodies are in clinical development for cancer (**Figure 2**, **Appendix Table S9**).

### Exploring associations of prioritised proteins with HCM-relevant cardiac outcomes

For the subset of five prioritised proteins, we subsequently explored potential pleiotropic effects on the HCM-relevant outcomes HF, DCM, SCA, and VA. NCAM1 was the only risk-decreasing prioritised protein and NCAM1 was only associated with HF. The other prioritised proteins associated with all four relevant outcomes. The effect direction were concordant to their association with HCM, CMP, or LVMWT (**Figure 4**, **Appendix Table S10**).

**Figure 4.**
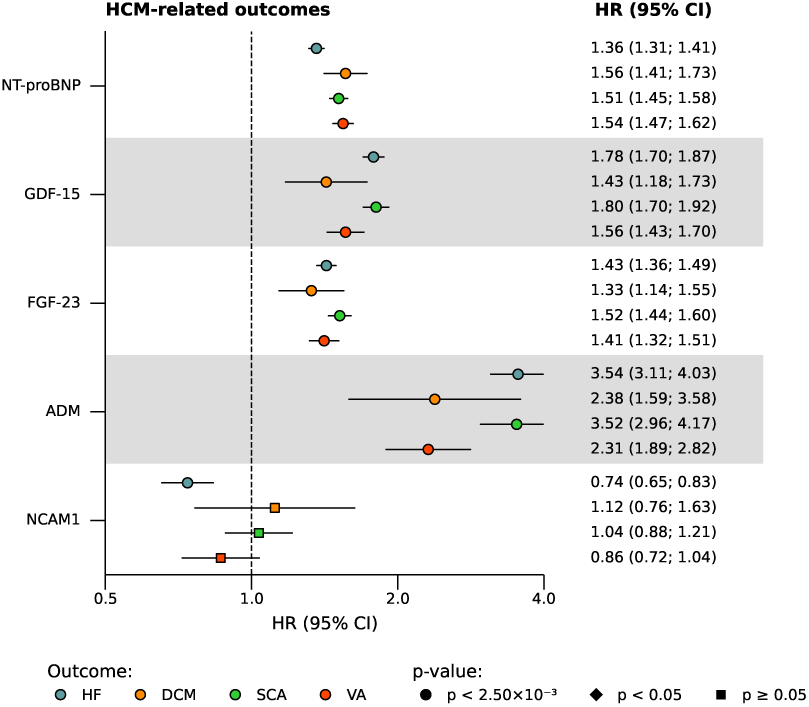
Effect estimates for the associations of the prioritised proteins with HCM-relevant outcomes. Proteins were considered significant if the p-value was below the p-value threshold of 0.05 divided by the number of proteins (8) and outcomes (4) tested. Abbreviations: CI = confidence interval; DCM = dilated cardiomyopathy; HCM = hypertrophic cardiomyopathy; HF = heart failure; MD = mean difference; SCA = sudden cardiac arrest; VA = ventricular arrhythmias.

## Discussion

The current study has identified and prospectively replicated 21 plasma proteins for their involvement in HCM progression and onset. Following a multi-tiered approach, initial identification was based on disease severity in a cohort of truncating *MYBPC3* pathogenic variant carriers and replication was based on associations with incident HCM or CMP and CMR derived LVMWT in a general population cohort. Five of these proteins (NT-proBNP, GDF-15, FGF-23, ADM, and NCAM1) were prioritised based on cardiac tissue expression and druggability, highlighting their potential as markers for diagnostics, disease monitoring, and drug targets.

Previous studies have identified several candidate biomarkers for HCM^17,18^ and primarily focused on distinguishing HCM patients from controls with hypertensive left ventricular hypertrophy or other cardiomyopathies^19–21^. This implicated the Ras-MAPK pathway in HCM^19–21^, in which two of the prioritised proteins from this study (FGF-23 and NCAM1) are involved. Our discovery cohort uniquely included only *MYBPC3* truncating pathogenic variant carriers and captured the full clinical HCM spectrum at the time of protein measurements, ranging from unaffected pathogenic variant carriers to those with advanced disease. This enabled the identification of proteins involved in both onset and progression of HCM. The replication of protein associations in the UK Biobank with adjustment for age and sex confirms that the prioritised proteins are age-independent and likely represent disease severity markers. Three of the prioritised proteins, NT-proBNP, FGF-23, and ADM, were previously identified as biomarkers in a sarcomeric HCM patient group^22^, further confirming our results. We subsequently leveraged the population cohort UKB to replicate the proteins in a general population study free of CMP diagnosis at the time of protein measurements and considering HCM and CMP diagnoses irrespective of genetic carrier status. While the replication step allowed for additional generalisation to non*-MYBPC3* carriers, it also implies that variant-specific or disease-state-specific proteins may have been missed. Replication and prioritisation were applied to ensure robust and independent findings, but absence of replication or prioritisation should not be overemphasized^23,24^ and untargeted approaches could yield additional insights.

NT-proBNP and GDF-15 were identified as important for all HCM severity comparisons indicating HCM onset (unaffected vs. affected, unaffected vs. mild, and unaffected vs. severe). Both proteins have previously been associated with HCM and HF^18,25–28^, and GDF-15 is proposed as a potential HF drug target^29^, supporting the validity of our findings.

Additionally, ADM, FGF-23, and NCAM1 were implicated in two HCM severity comparisons indicating HCM onset and replicated in the UKB, where they were robustly associated with HCM-relevant outcomes. Each of these proteins is targeted by approved or developmental drugs and therefore provide potential opportunities for drug repurposing. ADM a vasodilative protein associated with the renin-angiotensin-aldosterone pathway, which is involved in hypertrophy and fibrosis^30^. While ADM is indirectly affected by drugs targeting the RAAS-pathway, such as ACE inhibitors and angiotensin receptor blockers, it can also be directly targeted by enibarcimab, which is postulated to prevent congestion in HF through enhanced vascular integrity^31^. Targeting ADM may help prevent downstream myocardial remodelling and reduce afterload through its vasodilatory actions. Together, these properties suggest ADM may be a useful target in HCM as well, irrespective of genotype. FGF-23 is a hormone associated with HF and mortality^32,33^ and is implicated in LV hypertrophy^34,35^. It is inhibited by burosumab to treat X-linked hypophosphatemia. We found FGF-23 to be overexpressed in the heart and its association with HCM suggests a pathogenic role, supporting further investigation of FGF-23 inhibition as a potential disease-modifying strategy in HCM. A genome-wide association study identified genetic variants in *NCAM1* associated with LVMWT^36^, supporting our findings. NCAM1 is upregulated in stressed cardiomyocytes, where it is associated with reduced apoptosis^37,38^. It was also proposed as an HF drug target due to its disruptive effect on cardiomyocyte to cardiomyocyte adhesion, impairing electrical conduction and contractile function^39^. NCAM1 is targeted by antibodies that are in development to treat cancers, which could be explored for potential repurposing in HCM.

This study provides a first step in identifying protein biomarkers useful for monitoring HCM progression and opportunities for improving patient care. The discovery of these proteins in *MYBPC3* pathogenic variant carriers indicates they may aid in early identification of variant carriers at risk of disease development and progression, complementing imaging and genetic diagnostics. In addition to the drug repurposing opportunities, integrating the five identified biomarkers into routine clinical evaluation may potentially enhance risk stratification. The proteins are implicated in important pathophysiological processes such as fibrosis and hypertrophy, indicating they may serve as biomarkers to evaluate disease progression and treatment response. Future studies using independent data can provide further evidence by evaluating longitudinal protein patterns to assess their utility for guiding clinical practice.

The discovery cohort contains only carriers of truncating *MYBPC3* variants and is therefore genetically homogeneous, thus aiding the identification of proteins important in both disease onset as well as progression. Generalisability to HCM patients with non-*MYBPC3* truncating variants or no genetic variants was explored through replication in the unslected UKB sample, supporting a broader relevance of the replicated proteins beyond *MYBPC3*-related HCM. However, clinical phenotypes in the UKB rely on ICD10-codes, which may include only the patients in later disease stages and is less specific, as patients can also be coded under broader terms, such as CMP. Nevertheless, focusing on the proteins consistently associated with HCM, CMP, or LVMWT across the discovery cohort and the UKB provides a set of biomarkers with strong potential for broader clinical relevance.

We have identified and replicated 21 plasma proteins associating with disease progression in *MYBPC3* pathogenic variant carriers, as well as with onset of disease and LVMWT in unselected HCM. These findings therefore prioritise proteins that associate with HCM independent of genotype and provide leads for disease monitoring and drug targets.

## Data Availability

Both data and samples for BIO FOr CARE are available to external researchers through submission of an application to our data access board, which consists of investigators from each participating centre.

## Non-standard abbreviations and acronyms

CI: confidence interval
CMP: cardiomyopathy
CMR: cardiac MRI
DCM: dilated cardiomyopathy
HCM: hypertrophic cardiomyopathy
HF: heart failure
HR: hazard ratio
LV: left ventricular
LVMWT: LV maximum wall thickness
SCA: sudden cardiac arrest
UKB: UK Biobank
VA: ventricular arrhythmia

## Funding

MvV is supported by the postdoc talent grant from Amsterdam Cardiovascular Sciences. AFS is supported by the UK Research and Innovation (UKRI) under the UK government’s Horizon Europe funding guarantee EP/ Z000211/1, BHF grant PG/22/10989, the UCL BHF Research Accelerator AA/18/6/34223, the UCL BHF Centre of Research Excellence RE/24/130013. This publication is part of the project “Computational medicine for cardiac disease” with file number 2025.027 of the research programme “Computing Time on National Computer Facilities” which is (partly) financed by the Dutch Research Council (NWO). This work was supported by the Netherlands Cardiovascular Research Initiative with the support of the Dutch Heart Foundation (grant Dutch Cardiovascular Alliance 2020B005 DoubleDose), the National Institute for Health Research University College London Hospitals Biomedical Research Centre, and an unrestricted grant from Bristol Meyers Squibb. ATR is funded by ZonMW grant (2024 Clinical Fellows; grant no. 09032232310042) and HORIZON cardiogenomics pathfinder IMPACT (grant no. 101115536).

## Disclosures

AFS has received funding from New Amsterdam Pharma for unrelated projects.

## Acknowledgements

This research has been conducted using the UK Biobank Resource under Application Number 24711 and 12113.

## Notes

### Author Declarations

This study was performed in accordance with the Helsinki declaration and was approved by the Medical Ethics Committee of the UMC Utrecht under registration number NL5588904115.

