## Appendix for "Identifying and replicating plasma proteins associated with hypertrophic cardiomyopathy severity in carriers of pathogenic *MYBPC3* variants"

Hassanzada *et al.*

### Appendix

#### Figure legends

**Figure S1. Overview of the 27 proteins associated with HCM severity and their overlap in the HCM severity contrasts**

**Figure S2. Percentage of mRNA expression across tissues**

For visualisation purposes, only tissues where at least one protein was overexpressed are depicted. Proteins are ordered by cardiac expression and tissue overexpression is indicated by diagonal stripes.

**Figure S3. Top 20 Reactome pathways among the prioritised proteins**

The dotted line indicates an adjusted $-\log_{10}$ p-value threshold of 0.05. Pathways with a $-\log_{10}$ p-value larger than that are considered significantly enriched.

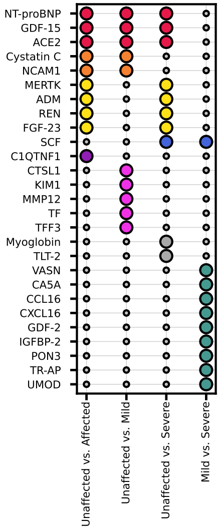
NT-proBNP
GDF-15
ACE2
Cystatin C
NCAM1
MERTK
ADM
REN
FGF-23
SCF
C1QTNF1
CTSL1
KIM1
MMP12
TF
TFF3
Myoglobin
TLT-2
VASN
CA5A
CCL16
CXCL16
GDF-2
IGFBP-2
PON3
TR-AP
UMOD
Unaffected vs. Affected
Unaffected vs. Mild
Unaffected vs. Severe
Mild vs. Severe

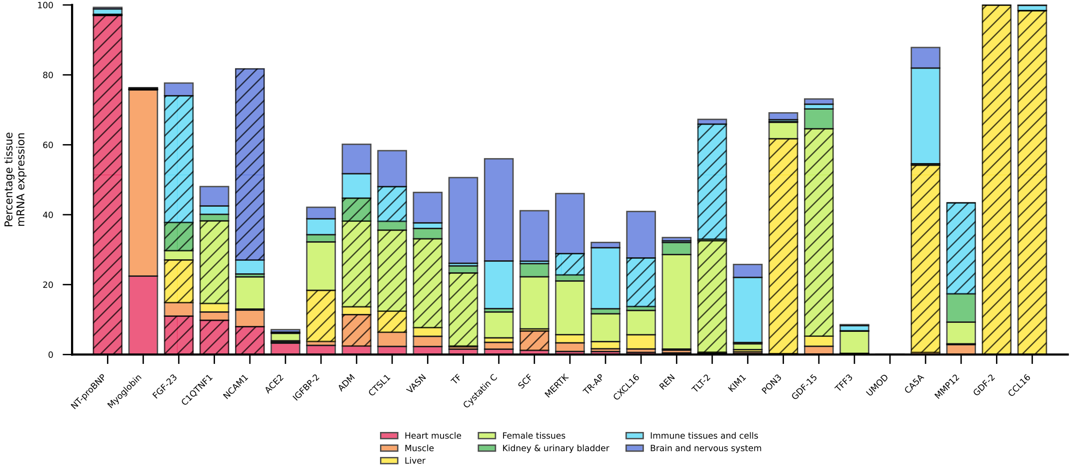

Percentage tissue mRNA expression
0
20
40
60
80
100
NT-proBNP
Myoglobin
FGF-23
C1QTNF1
NCAM1
ACE2
IGFBP-2
ADM
CTSL1
VASN
TF
Cystatin C
SCF
MERTK
TR-AP
CXCL16
REN
TLT-2
KIM1
PON3
GDF-15
TFF3
UMOD
CA5A
MMP12
GDF-2
CCL16
Heart muscle
Muscle
Liver
Female tissues
Kidney & urinary bladder
Immune tissues and cells
Brain and nervous system

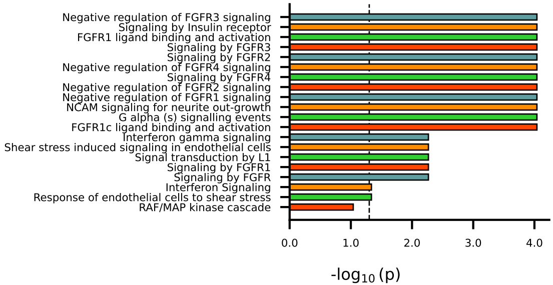

Negative regulation of FGFR3 signaling
Signaling by Insulin receptor
FGFR1 ligand binding and activation
Signaling by FGFR3
Signaling by FGFR2
Negative regulation of FGFR4 signaling
Signaling by FGFR4
Negative regulation of FGFR2 signaling
Negative regulation of FGFR1 signaling
NCAM signaling for neurite out-growth
G alpha (s) signalling events
FGFR1c ligand binding and activation
Interferon gamma signaling
Shear stress induced signaling in endothelial cells
Signal transduction by L1
Signaling by FGFR1
Signaling by FGFR
Interferon Signaling
Response of endothelial cells to shear stress
RAF/MAP kinase cascade
0.0
1.0
2.0
3.0
4.0
-log10 (p)
